# Genomic Machine Learning Meta-regression: Insights on Associations of Study Features with Reported Model Performance

**DOI:** 10.1101/2022.01.10.22268751

**Authors:** Eric Barnett, Daniel Onete, Asif Salekin, Stephen V Faraone

**Affiliations:** Department of Neuroscience and Physiology, SUNY Upstate Medical University, Syracuse, New York, USA; College of Medicine, MD Program, SUNY Upstate Medical University, Syracuse, New York, USA; Syracuse University; Department of Psychiatry and Behavioral Sciences, SUNY Upstate Medical University, Syracuse, New York, USA

## Abstract

**Background:** Many studies have been conducted with the goal of correctly predicting diagnostic status of a disorder using the combination of genetic data and machine learning. The methods of these studies often differ drastically. It is often hard to judge which components of a study led to better results and whether better reported results represent a true improvement or an uncorrected bias inflating performance.

**Methods:** In this systematic review, we extracted information about the methods used and other differentiating features in genomic machine learning models. We used the extracted features in mixed-effects linear regression models predicting model performance. We tested for univariate and multivariate associations as well as interactions between features.

**Results:** In univariate models the number of hyperparameter optimizations reported and data leakage due to feature selection were significantly associated with an increase in reported model performance. In our multivariate model, the number of hyperparameter optimizations, data leakage due to feature selection, and training size were significantly associated with an increase in reported model performance. The interaction between number of hyperparameter optimizations and training size as well as the interaction between data leakage due to optimization and training size were significantly associated reported model performance.

**Conclusions:** Our results suggest that methods susceptible to data leakage are prevalent among genomic machine learning research, which may result in inflated reported performance. The interactions of these features with training size suggest that if data leakage susceptible methods continue to be used, modelling efforts using larger data sets may result in unexpectedly lower results compared to smaller data sets. Best practice guidelines that promote the avoidance and recognition of data leakage may help the field advance and avoid biased results.

## Introduction

The genetic study of complex disorders has made great strides in the discovery of genome-wide significant genetic loci and substantial evidence for polygenicity[1]. These discoveries have generated new hypotheses about the etiology of these disorders and have motivated machine learning (ML) efforts to separate cases and controls using genome-wide association study (GWAS) data. While results from early genomic ML research had been promising, the potential pitfalls of such studies have limited their interpretation[2]. Although best practices have previously been described, the methods, reporting, and overall study design for genomic ML studies vary so drastically that it is often difficult to compare and evaluate studies[3]. This between-study heterogeneity may contribute to distrust and underutilization of machine learning results. A clearer understanding of which genomic ML research are most important in producing successful results could lead to more consistent study design and reporting, which may collectively move the field towards using genomic ML models as a component of personalized medicine.

To better appreciate the strengths and weaknesses of genomic ML research, one must understand the differences between ML analyses and traditional GWAS. GWAS seeks to determine loci that are statistically significantly different between cases of a disorder and healthy controls and to test for polygenicity[4]. Since these studies examine hundreds of thousands to millions of loci, researchers apply stringent genome-wide significance thresholds (most commonly p < 5 × 10^−8^) to reduce reporting false positive results.

In the ML analyses we review here, the primary goal is accurately predicting whether subjects are cases or controls using differences in loci between the two groups. Towards achieving this goal, relying only on loci that meet the threshold for genome-wide significance limits the learning capability of ML models. For example, a schizophrenia GWAS found that while 108 genome wide significant loci were able to explain 3.4% of the variation on the liability scale, including loci that met the nominal significance threshold (0.05) increased the variation explained to 7%[5]. This effect may be more pronounced in ML models which take advantage of interactions between loci to find patterns that are useful in differentiating cases and controls since more loci give the models more potential to find patterns.

Including additional loci in ML models has some drawbacks. One of the most important aspects of generalizable ML models is avoiding overfitting, which becomes more difficult as the number of loci increases[6]. Overfitting occurs when a model learns patterns that are only present within the data used to train the model (See Figure 1). In ML algorithms, the training process learns model parameters that minimize the difference between the predicted and actual case/control labels. Researchers aim to build models that use real, generalizable differences between cases and controls to make each prediction, but in practice, models are free to use whatever patterns best minimize that difference. If a model memorizes the noise specific to only the training data, the model is less motivated to learn patterns that may be more generalizable if using those patterns is less successful than memorizing training data noise. For many ML models, constraints are added to reduce the model’s ability to overfit, but overfitting is rarely completely avoided[6]. Each additional locus that a model has access to increases the probability of overfitting but also has the potential to add generalizable information the model can use to increase its ability to separate cases and controls.

**Figure 1:**
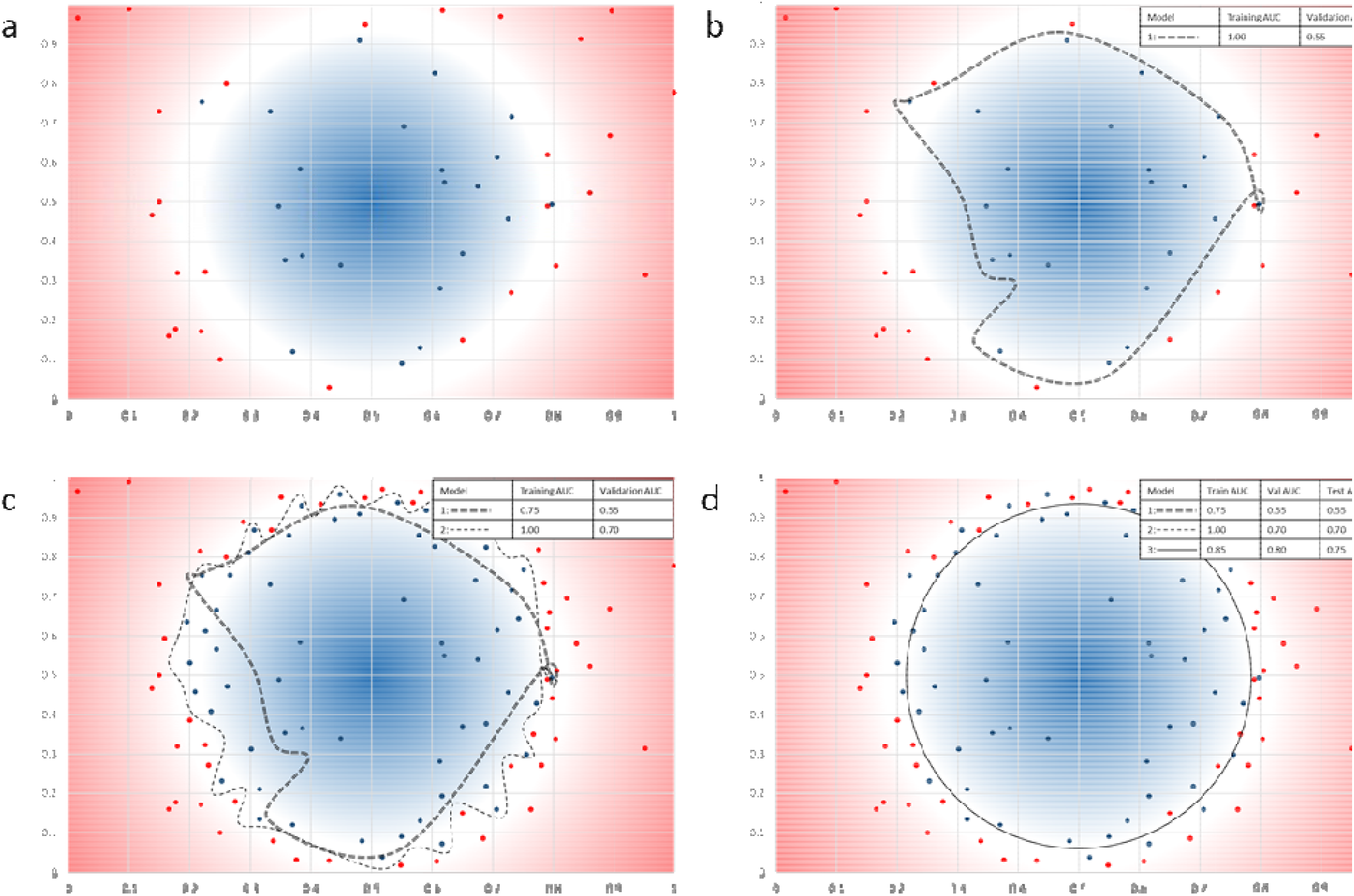
Overfitting, sample size, and data leakage a. **Plot Overview:** In this illustrative example, we have condensed most of the known genetic variance for a hypothetical disorder into 2 continuous features scaled from 0 to 1. In these graphs the axes represent those 2 continuous features, and each point represents a person in the training subset based on their values for each feature. People who are cases of the hypothetical disorder are shown in red while people who are controls are shown in blue. The true population distribution of cases and controls, which would be unknown to us when modeling since we can only sample from the population, is represented by the background, with the intensity of the gradient representing how likely a person is to be a case (red) or control (blue) given the 2 features. The white space represents a region in which the likelihood of being a case or control is similar and perfect separation of cases and controls is impossible with only genetic data. Our task is to build a model that predicts whether each person is a case or control. To do this task in this example, we will build models that enclose all predicted controls. The optimal solution in this case would be a circle splitting the white region, but since our data are not a perfect representation of the population distribution, the models we make will inevitably be less optimal. We will discuss several factors that commonly impact how similar a model can be to the optimal model. b. **Overfitting:** Many models are easily capable of achieving perfect prediction on the data that are used to build or train the model. However, that performance may be specific to the training data, as illustrated by the overfit Model 1. To get a more generalizable measure of model performance, we should test performance in data that are not used to train the model. If we were to test Model 1 in a different sample of unseen data (validation subset) we may find that it is not nearly as predictive in that subset since the model was trained using some patterns that generalize to the population and some patterns in the training data that do not match the population distribution. c. **Advantages of Increasing Training Size:** If we increased the training size (only case/control boundary examples illustrated) it would become clear that Model 1 was overfit to the original data and the model does not generalize to the population. At this point it may still be possible to get perfect prediction by overfitting the model to the data with more complex models, as illustrated by Model 2, but overfitting becomes increasingly difficult as more data are added to the training subset. d. **Bias due to Data Leakage:** To find the model with the best prediction in unseen data, we can use a validation subset (not shown) to optimize which model type we use and which hyperparameters or options to use within a model. During this process, we indirectly learn information about the validation subset as we find the model that best predicts that specific set of unseen data. Therefore, our model is biased towards correctly predicting the validation subset due to indirectly “leaking” information from the validation subset during model optimization. Each additional model trained during optimization increases the potential for data leakage, but also increases the potential of finding the optimal model. The best model among many trials optimizing towards the validation subset is illustrated by Model 3. Since the data in the validation subset are no longer truly unseen data, we should test the best model in a new, completely unseen subset to avoid data leakage from inflating our reported model performance. We can use the test subset (not shown) for this purpose to report the performance of the optimized model on unseen data. In this example, the validation data contained more cases than controls in the unshaded area wherein the population is equally split between cases and controls. This resulted in choosing a model that performs better in the validation subset compared to the test subset because the test subset and population do not have all the same patterns seen in the validation data. The imperfect generalization of models to unseen data is expected, which is why a final test on truly unseen data is important for accurate performance reporting. Testing more models on the test subset may lead to the same indirect data leakage occurring in the test subset, resulting in inflated reported performance, so use of the test subset should be reserved for the final model.

Many ML researchers account for overfitting by testing the performance of their models on data that were not seen during training[7]. One of two methods is typically used: cross-validation within the training set and validation with data not used at all in the training process. In k-fold cross-validation, researchers randomly split the data into some number of subsets, called folds, and then complete the association analysis and modeling that detect and use loci that are different between cases and controls to best separate the two classes using all but one of the folds. The model is trained using data from k-1 folds and its accuracy is tested in the single fold that was not included in training. This process is repeated until the model has used each fold as the withheld subset. Then the results across all iterations are averaged.

In the hold-out method, researchers randomly split the data into either two (training and test) or three (training, validation, and test) subsets. The training subset is used for the association analysis and modeling. If present, the validation subset is used to tune the model by setting the optimal hyperparameters, which are all the options and configurations that are not trained by the model itself, to best predict the validation subset. Then, the test subset is used to measure and report model performance. Unlike in cross-validation where all folds are the same size, the hold-out method typically uses 60-80% of the data in the training subset, while the remaining data is split evenly between the validation and test subsets. Cross-validation is often used when data is limited since in this method the model has a chance to train using each person in the study. The hold-out method, while not allowing the model to train on each person, is thought to be the more conservative approach and less likely to produce an overfit model.

The value of external testing can easily be lost through methods that leak information about the test subset/fold into the training of a model[8]. When this occurs, the model can use that information to model the specific test data more accurately. Consequently, the test data no longer represent unseen data and no longer account for overfitting to the same degree. The result in a model that is biased in favor of the test data. This problem, called data leakage, is especially detrimental because it often goes unnoticed, leading researchers and their audiences to believe that their model performs exceptionally well even outside the training subset when instead they are observing model bias. The amount of data leakage caused through methodological issues can vary from slight leakages that may result in some overreporting of performance, to major leakages that cause the testing subset/fold to mimic the training performance with near-perfect prediction. One example of minor data leakage could be a cycle of checking the performance of a model in the test subset, then deciding to try more types of models using the same data subsets until the performance meets a threshold deemed worthy of reporting. In this situation, researchers indirectly leak information about the test data into their modeling, allowing them to select the model that performed best in the test data and reporting an inflated result (See Figure 1). An example of major data leakage is using the entire data set to select which of the many different features best separate cases and controls and only splitting the data into subsets during the actual modeling. In this situation, when the features were selected with the entire data set, information about the test data were directly leaked into the process and the features included in the model will perform well in the test data, but in unseen data will either be less predictive or entirely unpredictive if the features selected are specific to the data in the feature selection process.

The choices researchers make regarding which ML models to train, loci to include, and methods to use for measuring performance and optimization are critical decisions that will determine the outcome and validity of their study. But since few studies compare ML variations in the same external data sets, comparisons between studies are difficult even within the same disorder. This leads to a potential dilemma: Is a model with a higher reported performance better than a lower performing model or more overfit to their data?

Here, we report a systematic review that extracted information on model performance, disorder, training size, ML methods, optimization methods, performance measurement methods, and reporting on common issues from all published genomic ML papers. We use this information in a mixed-effects linear regression model predicting model accuracy as measured by the area under the receiver operating characteristic curve (AUC). Within each data set and study involving machine learning models, many factors, such as training size, model type, and disorder, likely have a real, generalizable impact on reported model performance. When looking at a group of studies, those factors may be harder to identify due to the variable effects of data leakage among the studies in the group. We hypothesized that data leakage would be associated with a significant increase in reported AUC. We sought to test this hypothesis and to identify other study features that lead to increased reported AUC.

## Methods

### Search strategy and selection

To identify studies that used genotype data as input for machine learning models to predict any disorder, we searched PubMed using the key-words ‘(GWAS OR genotype[ti] OR SNP) AND (classification OR predict OR prediction) AND (“machine learning” OR “data mining[ti]” OR “neural network*” OR “random forest” OR “support vector machine”)’. The search produced 660 studies (up to July 1, 2021).

We excluded studies that were not using machine learning classifiers to predict a human disorder, studies that did not report results for genotype only models, and studies that did not report testing performance outside the data used to train the model. We also excluded studies starting with less than 100 variants, since these studies are generally focused on specific variants that have been thoroughly studied and do not face many of the challenges addressed in this paper. We excluded studies that did not report AUC as a performance metric since combining different performance metrics in our analysis would limit interpretation and potentially bias results. After exclusion criteria, 41 studies remained[9-49]. These studies provided accuracy statistics for 77 models because some studies modeled multiple disorders. Supplementary Figure 1 shows the article selection procedure in a PRISMA diagram.

### Data extraction

We extracted the following data from each included study: disorder predicted, number of subjects used for training and testing, number of participants the model was trained on, highest AUC, method for testing/reporting model AUC, reporting on optimization, number of hyperparameters optimized, number of models reported, and machine learning method with the highest AUC. We split studies that modeled multiple disorders such that each row of extracted data represented a single disorder from a single study.

### Regression analysis

We fit linear mixed-effects models to test the individual and combined contribution of study variables to AUC. The standard error of each of our models was estimated using a clustered sandwich estimator clustering on PMID of the included studies, which adjusts standard errors for the lack of statistical independence of results within studies. We fit univariate models with AUC as the dependent variable and used the following as independent variables: data leakage through feature selection, data leakage through hyperparameter optimization, disorder type, reporting of optimization, number of hyperparameter optimizations reported, disorder heritability, model type, testing method, and size of training dataset.

Data leakage through feature selection was a binary feature scored as 1 if the data used to test model performance were used to select which features would be included in the model. It was scored zero otherwise. Data leakage through optimization was a binary feature scored 1 if the data used to test model performance was used at all in optimizing the model. It was scored zero otherwise. Reporting of hyperparameter optimization was a binary variable scored 1 if the authors reported any optimization of model hyperparameters. It was scored zero otherwise. Number of hyperparameters optimizations indicated the number of hyperparameters the authors reporting optimizing in the model with the highest AUC. Disorder heritability for each disorder included in these studies was gathered from heritability studies of those disorders based on twins or families. Model type was a binary variable scored 1 if the model with the highest AUC was non-linear and 0 if it was linear. Testing method was a binary variable scored 1 if cross-validation was used to test model performance and 0 if a hold-out test subset was used to test model performance.

We corrected univariate p-values for multiple testing using Bonferroni correction. Variables were added to a multivariate model sequentially, ordered based on the p-value of the variables univariate model, and kept if the variable remained significant.

Among the possible interaction terms for the variables used in our models, we identified 6 interactions that could be reasonably hypothesized to impact prediction performance. These 6 terms were: data leakage through feature selection + training size, number of hyperparameter optimizations + training size, testing method + training size, data leakage through hyperparameter optimization + training size, number of hyperparameter optimizations + data leakage through hyperparameter optimization, and number of hyperparameter optimizations + testing method. We tested the potential interaction terms as individual additions to the multivariate model, added the interactions sequentially ordered by the p-value of the initial interaction models, and kept the interactions in the final model if they remained significant. For each significant interaction, we estimated predictive margins using STATA16’s margins command, which computes the average probability for each observation at a fixed level of a selected variable[50]. In our analyses, these predictive margins estimate the average AUC of our collection of studies at each fixed level of a selected variable. To visualize these effects within the training size range of the included studies, we fixed the values of tested variables at each level, estimated the predicted AUC for each study, and then plotted a linear prediction of the predicted AUCs for each group level as a function of training size.

## Results

Among the 41 studies and 77 models included, 27 different disorders were modeled. The average AUC among all models was 0.77. Thirty-nine percent (N = 30) of the models modeled eight autoimmune disorders and had a mean AUC of 0.81 ± 0.12. Eighteen percent (N = 14) of the models modeled six psychiatric disorders and had a mean AUC of 0.74 ± 0.16. The remaining 33 models modeled 12 disorders that did not fit into either of these groups and had a mean AUC of 0.76 ± 0.16. The included studies and models are listed in Table S1.

In univariate models the number of hyperparameter optimizations reported and data leakage due to feature selection were significantly associated with an increase in AUC in the test data after correcting for multiple testing (Table 1). Use of a non-linear model was nominally associated with an increase in AUC but did not remain significant after correcting for multiple testing. Disorder type, reporting on optimization, training size, testing method, disorder heritability, and data leakage due to optimization were not significant.

**Table 1.** Univariate mixed-effects linear model results

| Feature | Coefficient | z | P>\|z\| |
| --- | --- | --- | --- |
| Number of optimizations | 0.02 | 7.07 | <0.001** |
| Data leakage: feature selection | 0.17 | 3.65 | <0.001** |
| Model type | 0.12 | 2.63 | 0.008 |
| Disorder type |  | 1.95 | 0.15 |
| autoimmune vs other | -0.05 | -1.51 | 0.14 |
| autoimmune vs psych | -0.06 | -1.63 | 0.09 |
| psych vs other |  | 0.49 | 0.63 |
| Reported optimization | 0.06 | 1.57 | 0.12 |
| Training size | $-7 \times 10^{-7}$ | -0.74 | 0.46 |
| Testing method | 0.04 | 0.73 | 0.46 |
| Disorder heritability | 0.07 | 0.61 | 0.54 |
| Data leakage: optimization | 0.03 | 0.56 | 0.57 |

\*p<0.05 after multiple testing correction, \*\*p<0.01 after multiple testing correction

When interactions were entered one at a time, the interaction between number of hyperparameter optimizations and training size as well as the interaction between data leakage due to optimization and training size were significantly associated with AUC after correcting for multiple testing (Table 2). In predictive margins analysis, the average marginal effect of training size on AUC was nominally significantly positive (p = 0.004) when data leakage due to hyperparameter optimization was absent and was not significant (p = 0.71) when data leakage due to optimization was present. Figure 2 illustrates the effect of predictive margins analysis by showing the linear fit of the predicted AUC when we fixed data leakage due to hyperparameter optimization to either absent or present. The average marginal effect of training size on AUC was significantly positive (p < 0.001) when the number of hyperparameters optimized was fixed at zero and was significantly negative (p = 0.001) when the number of hyperparameters optimized was one standard deviation higher than the mean (Figure 3). After we added data leakage due to hyperparameter optimization to the previous predictive margins analysis, the marginal effect of training size on AUC with no hyperparameter optimization remained significantly positive (p = 0.001) while the marginal effect of training size on AUC with the number of hyperparameters optimized fixed at one standard deviation higher than the mean remained significantly negative (p = 0.002). Figure 3 illustrates the effect of this predictive margins analysis by showing the linear fit of the predicted AUC when we fixed number of hyperparameters optimized to either zero, which was approximately one standard deviation below the mean, and 6.6, which was approximately one standard deviation above the mean. When sequentially adding the interactions, only the interaction between number of hyperparameter optimizations and training size remained significant after Bonferroni correction.

**Table 2.**
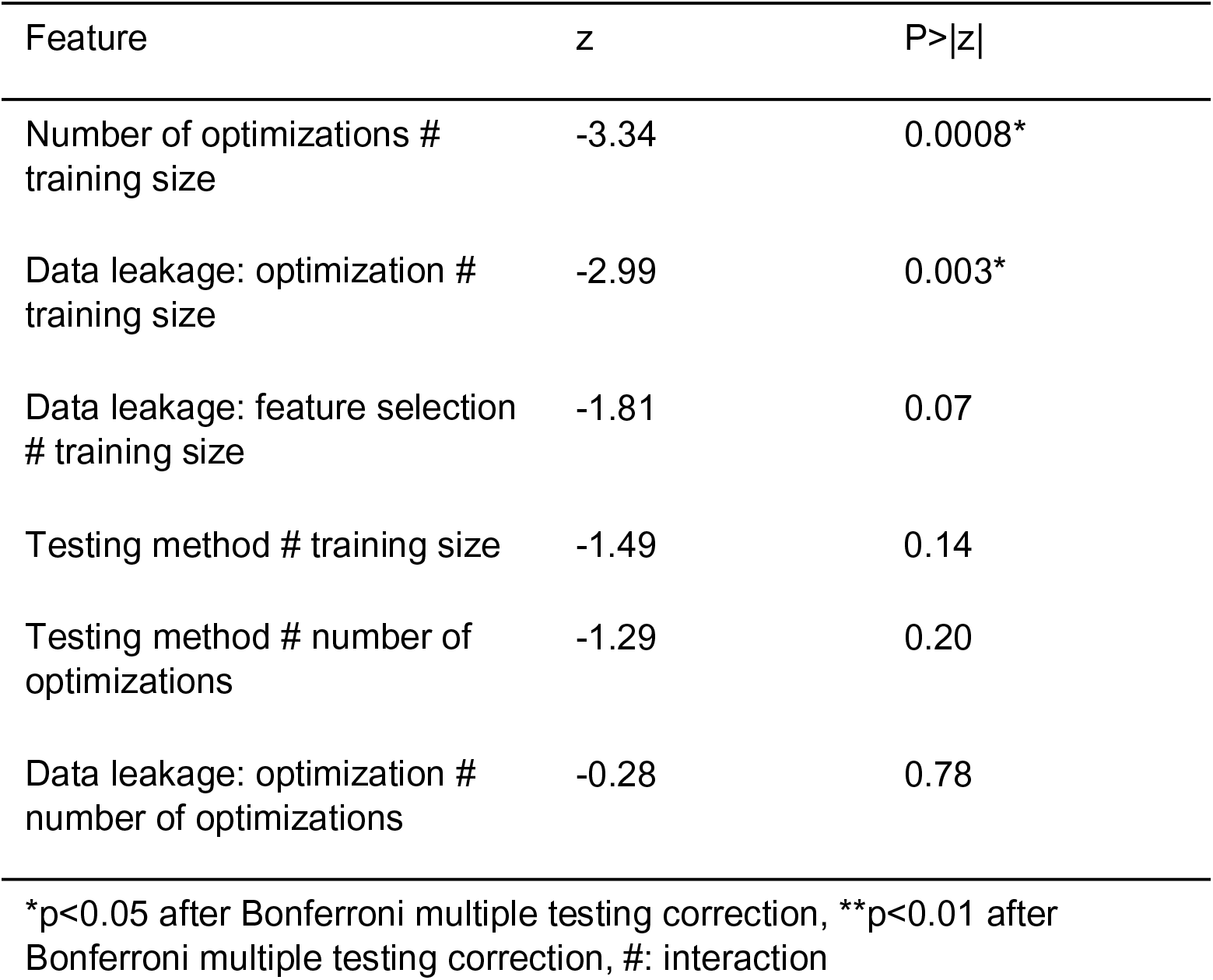
Multivariate mixed-effects linear model interaction results

**Figure 2.**
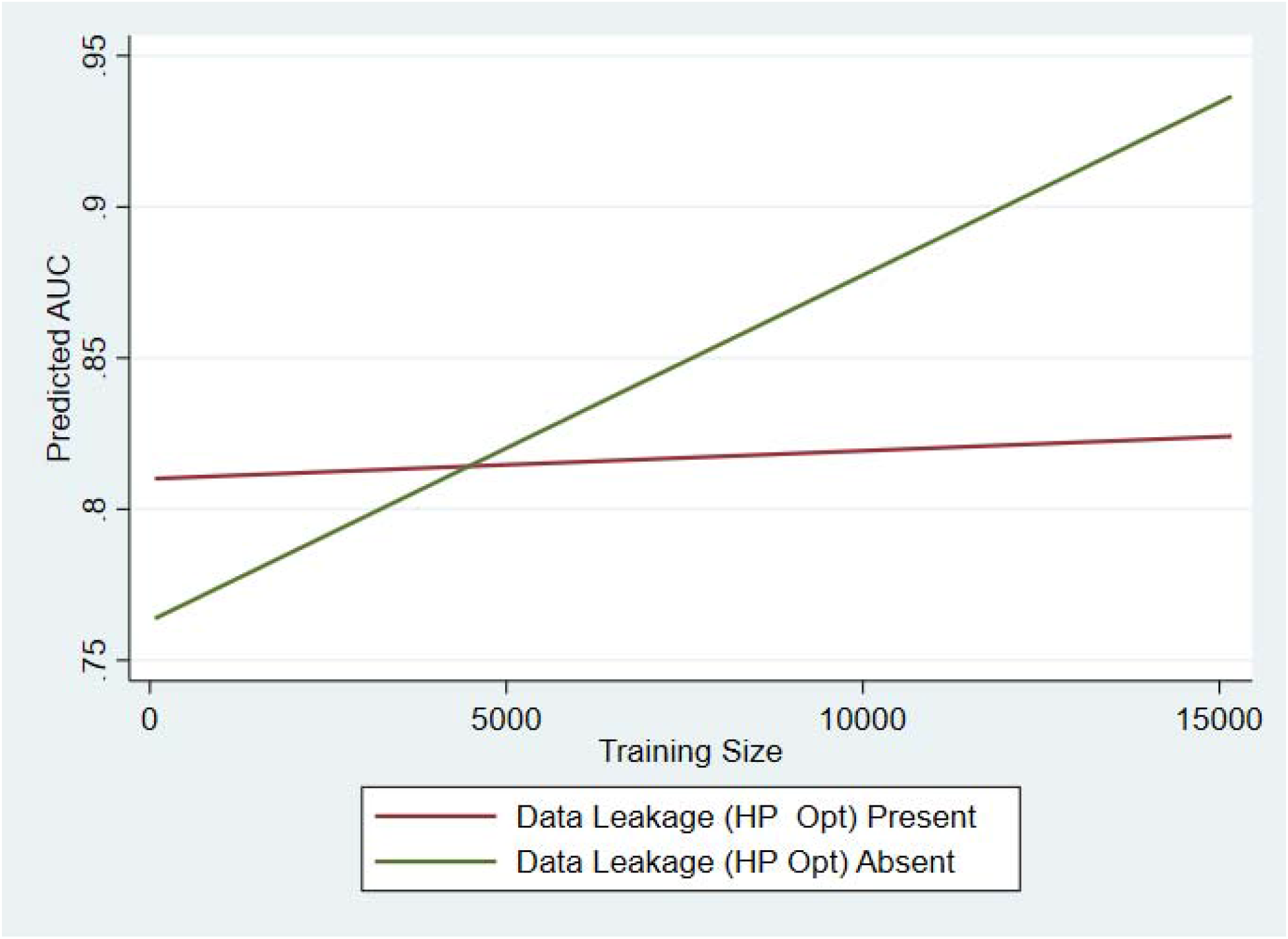
Predictive Margins Analysis of Data Leakage Due to Hyperparameter Optimization. Here, we graph the linear fit of the predicted AUC when we fixed data leakage due to hyperparameter optimization to either absent or present. The average marginal effect of training size on AUC was nominally significantly positive (p = 0.004) when data leakage due to hyperparameter optimization was absent and was not significant (p = 0.71) when data leakage due to optimization was present.

**Figure 3.**
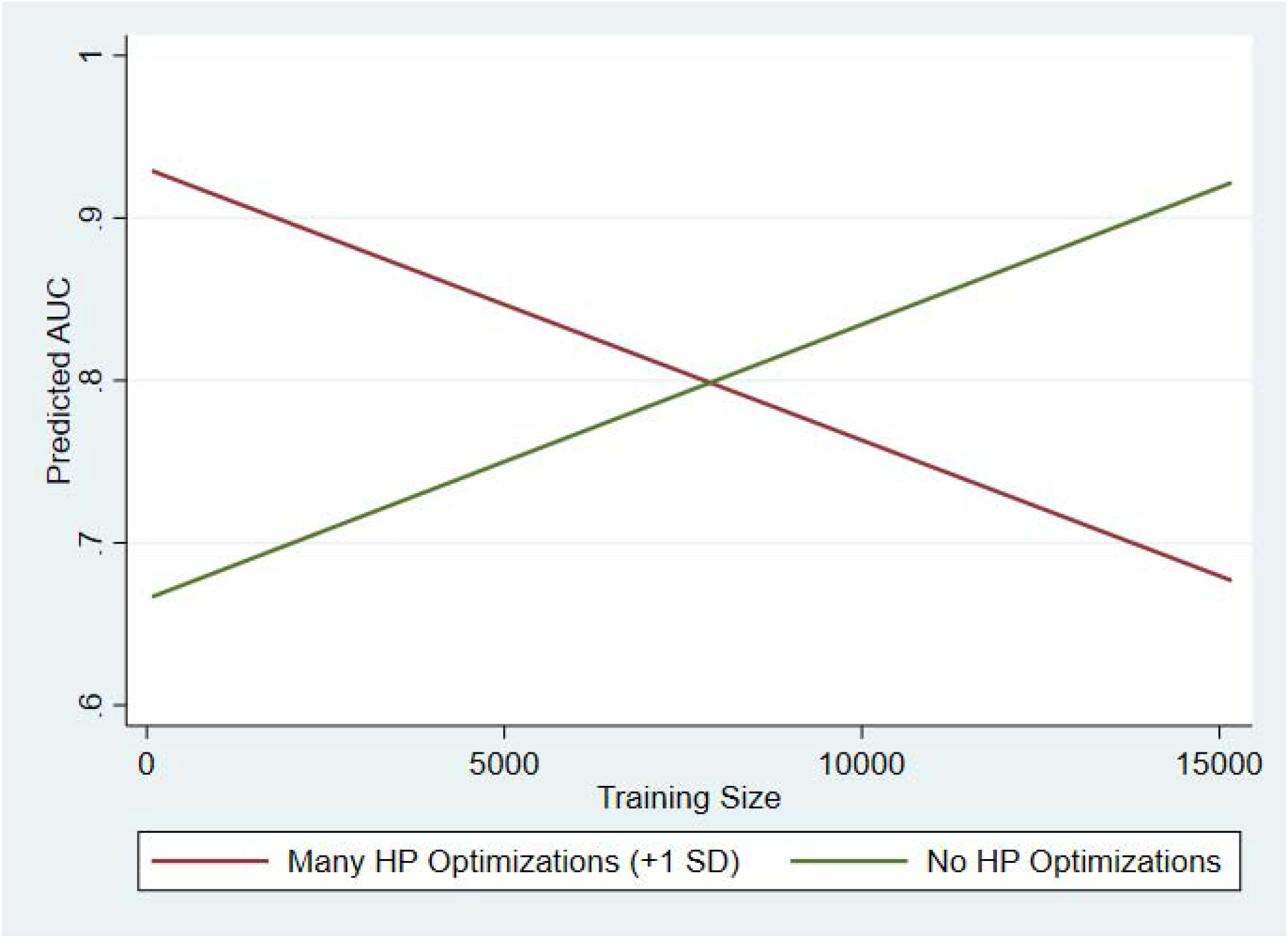
Predictive Margins Analysis of Number of Hyperparameters Optimized. Here, we graph the linear fit of the predicted AUC when we fixed the number of hyperparameter (HP) optimizations to either many optimizations (+1 standard deviations) or no hyperparameter optimizations. The average marginal effect of training size on AUC was significantly positive (p < 0.001) when the number of hyperparameters optimized was fixed at zero and was significantly negative (p = 0.001) when the number of hyperparameters optimized was one standard deviation higher than the mean.

Our final multivariate model included the number of hyperparameter optimizations, data leakage due to feature selection, disorder type, and the interaction between number of optimizations and training size along with their main effects (Table 3). In this model, number of hyperparameter optimizations, data leakage due to feature selection, and training size were significantly associated with an increase in AUC after correcting for multiple testing. In comparison to autoimmune disorders, both psychiatric disorders and disorders not belonging to either group were significantly associated with a decrease in AUC after multiple testing correction. The interaction between number of hyperparameter optimizations and training size was significantly negatively associated with AUC after correcting for multiple testing. The main effects of data leakage due to hyperparameter optimization, model type, reporting on optimization, testing method, and disorder heritability were not associated with changes in AUC in our multivariate model. The interaction between data leakage due to optimization and training size was initially significantly negatively associated with AUC (p = 0.0039) but was no longer significant after multiple testing correction. The interaction between data leakage due to optimization and the number of hyperparameter optimizations was initially significantly positively associated with AUC (p = 0.019) but was not significant after multiple testing correction. None of the other interaction terms we tested were significantly associated with AUC.

**Table 3.** Multivariate mixed-effects linear model results

| Feature | Coefficient | z | P>\|z\| |
| --- | --- | --- | --- |
| Number of optimizations | 0.04 | 4.04 | 0.0001** |
| Data leakage: feature selection | 0.16 | 3.19 | 0.0014* |
| Disorder type |  |  |  |
| autoimmune vs other | -0.10 | -4.27 | <0.0001** |
| autoimmune vs psych | -0.11 | -3.72 | 0.0002** |
| psych vs other |  | 0.64 | 0.51 |
| Training size | $1.4 \times 10^{-5}$ | 3.56 | 0.0004** |
| Number of optimizations #<br>training size | $-6.58 \times 10^{-6}$ | -3.34 | 0.0008** |

\*p<0.05 after Bonferroni multiple testing correction, \*\*p<0.01 after Bonferroni multiple testing correction, #: interaction

## Discussion

Our analysis to determine which features of genomic ML studies lead to significant increases in AUC found evidence that, in many studies, methodological issues result in overreporting of model performance. Out of the studies investigated here, 44% had some form of data leakage due to feature selection. The most common form of data leakage due to feature selection was using the entire dataset for the GWAS and later splitting the dataset into multiple subsets or folds. Since the test data were used to determine the loci that best separate cases from controls before ML modeling, even if researchers split the data into separate subsets during ML, information from the test data has already leaked into the process, which would lead to overestimates of accuracy. The coefficient of our multivariate model for this feature indicates that, on average, this form of data leakage increases the AUC by 0.18 after adjusting for other factors. For some applications, an increase in AUC of that magnitude would be enough to move models into a performance range that would falsely suggest clinical utility. Although this is the first demonstration of this problem for a collection of genomic ML studies, others have previously warned of the potential for this type of issue to interfere with results.

Out of the included studies, 56% had some form of data leakage due to hyperparameter optimization. In this form of data leakage, the test data are used multiple times as the researcher is optimizing the hyperparameters of a model to determine which set of those hyperparameters results in the best performance in the test data. A study using random training data showed that improper use of cross-validation, one example of data leakage due to hyperparameter optimization, leads to biased performance in the compromised dataset[51]. This type of data leakage allows the model to overfit the data by giving it many different opportunities to find a model that best separates those specific test data. It is typically used for smaller data sets where it is not feasible to create an independent test set. Instead, a more generalizable practice would be to either use a different subset or cross-validation within the training data to optimize hyperparameters and apply those hyperparameters to a single model in the test data[7].

Although the main effect of data leakage due to hyperparameter optimization was not significant, its interaction with training size was significant even after correcting for multiple testing. Learning curve analyses have demonstrated that increasing training size results in increased performance in machine learning since the model has more opportunities to learn patterns and features within the data[52, 53]. In learning curve analyses, a machine learning model is trained with an increasing number of training examples and prediction performance is measured at each training size[54]. Generally, models will gradually improve with training size until a plateau is reached. The amount of training data necessary to reach the plateau and the prediction performance at the plateau depend on the complexity of the model and the prediction task. Increased training size also makes it more difficult for ML models to find non-generalizable patterns within the noise of large datasets and the correlated increase in test size makes it more difficult to bias towards the test data when data leakage is present[55]. Thus, we thought that the interaction of data leakage due to hyperparameter optimization and training size would be important and included it in our analysis. This interaction was significant. As illustrated in Figure 2, training size was nominally positively associated with the predicted AUC only when data leakage due to hyperparameter optimization fixed at absent. This finding makes it evident that optimizing directly on test data should be avoided and verifies that increasing the number of subjects included in machine learning studies could further improve results. Our model also warns that if data leakage due to optimization occurs, studies with this issue may not benefit as much as expected from increasing training size.

The number of hyperparameter optimizations reported by researchers was associated with increased AUC in our univariate and multivariate models. Unlike the data leakage features, which are clearly a source of bias, the number of hyperparameters optimized could lead to genuine improvement, bias, or a balance of the two. The “no free lunch” theorem states that all optimization algorithms have the same performance when averaged over all possible tasks[56]. Applied in the context of machine learning hyperparameters, this means that it is impossible to know which type of model or hyperparameters within the model will perform best on a given dataset without prior knowledge. This maxim has led to the best practice of optimizing the hyperparameters of a model by either using a grid search over different values of those hyperparameters or using hyperparameter optimization algorithms and suggests that at least some of the improvement due to the number of hyperparameters optimized is due to genuine prediction improvement.

Conversely, the variable representing the number of hyperparameters optimized may also be a proxy of cryptic data leakage due to optimization. If data leakage due to hyperparameter optimization is present within the study, more optimizations will likely result in a model that is more biased in favor of the test data because the model has more opportunities to use the information it obtains about the test data through data leakage to find the optimal configuration for those data. This is similar to the data leakage illustrated in Figure 1, but instead of being only biased towards the unreported validation subset, the model would also be biased towards the test data. As illustrated in Figure 3, the average marginal effect between training size and AUC is significantly positive when fixed at no hyperparameters optimized and significantly negative when fixed at a value 1 standard deviation above the mean number of hyperparameters optimized. One explanation of this finding is that since the models with no hyperparameters optimized have fewer chances to use any leaked information about the test data and the results of increasing training size would represent the real improvements demonstrated in unbiased learning curve analyses. In comparison, if models with more optimized hyperparameters are more biased due to unaccounted data leakage, we would expect a more negative association with AUC when more hyperparameters are optimized because, in addition to the positive effects of training size, the increase in training and testing size will increase the difficulty of biasing towards the specific leaked test data.. To test whether these effects could be explained through the data leakage through hyperparameter optimization variable, we ran an additional predictive margins analysis which included the variable in the model. The results of the analysis were almost identical after including the data leakage through hyperparameter optimization variable, suggesting that the data leakage observed here is beyond what we could detect through reporting on methods. An alternate explanation could be that studies with highly optimized models, large training size, and relatively low performance share some other cryptic factor that drives the negative association of AUC with training size when hyperparameters optimized is fixed to a high value. While arguably a diminishing effect of training size on AUC with increased optimization may instead reflect the ability of larger datasets to overcome the limitations of unoptimized modeling better than smaller data sets, this argument would not explain the significantly negative average marginal effect of training size on AUC in models with more optimized hyperparameters. Unbiased model improvements due to increased optimizations are likely a component of the positive marginal effect on AUC found in our analysis, but any real effects are inseparable from the cryptic data leakage. Our results suggest that even when optimization is reportedly handled appropriately, data leakage due to hyperparameter optimization may still be present. These findings should not be interpreted as evidence that hyperparameter optimization should be avoided to reduce data leakage. Hyperparameter optimization is an important component of improving machine learning models and can be done in ways that avoid data leakage. Instead, our results points towards the necessity of establishing and following standard practices that are more careful in avoiding data leakage in the genetics machine learning field.

The type of disorder studied was significantly associated with AUC in our final multivariate model after Bonferroni correction. After adjusting for other significant predictors of AUC, models of autoimmune disorders had significantly higher AUCs compared to both psychiatric disorders and all other disorders. We were initially surprised by the lack of association between disorder heritability and AUC since disorders with a larger genetic component should theoretically be more predictable in genetic models, but the significance of disorder type may help explain disorder heritability’s lack of significance in our multivariate models. The hypothesis that disorder heritability may be associated with an increase in AUC assumes that the difficulty of extracting and modeling the genetic components is equivalent in all disorders. If the accessibility and ease of modelling genetic risk differ between disorders, we would no longer expect heritability to be associated with AUC. While this conclusion could previously be inferred by comparing the effect sizes of individual loci in GWASs of different disorders, our analysis provides further evidence that the genetic information from some groups of disorders may collectively differ in accessibility and ease of modelling. Differences in accessibility and ease of modelling could be due to differences in genetic architecture or differences in measurement (e.g., differences in misclassification rates).

Our study has several limitations that may have limited our ability to detect associations between the included features and AUC. The studies used in this analysis are heterogeneous due, in part, to the inclusion of any human disorder. These disorders likely have differing genetic complexities and optimal prediction performances, which may have limited our ability to detect differences in AUC based on the features of the study but also highlights the strength of the features that were detected despite this heterogeneity. We were unable to use specific disorders as a feature in our analysis due to the limited number of studies with each disorder. Excluding papers that did not use AUC as a performance metric limited the number of studies we could include in this analysis. Our study was also limited to using what was reported in these studies, which may have limited our ability to fully assess data leakage and optimization.

Our analysis of genomic machine learning studies has implications for defining best practices for genomic ML studies. Although such studies may eventually lead to clinically actionable risk calculators, publications that overestimate results will not be replicated, which could lead the field to prematurely abandon ML. We found data leakage due to both feature selection and optimization to be prevalent. The former leads to increased AUCs that likely overestimate the models’ true performance outside the training data while the latter limits or hides the effect of increasing training size. We also found evidence that suggests data leakage due to optimization may occur even when studies report methods that should minimize the effects of data leakage. If genomic machine learning methods and results are to be improved and trusted, researchers must recognize and avoid these issues. Thorough best practice guidelines that promote the avoidance of data leakage and other common issues will be critical as the field grows and advances.

## Data Availability

All data availability information and data used in this study can be found in the referenced primary GWASs.

## Acknowledgements

This project has received funding from the European Union Horizon 2020 research and innovation programme grant agreement No 667302. This project has received funding from the European Union’s Horizon 2020 research and innovation programme grant agreement No 965381.

## Data Availability

All data produced in the present study are available upon reasonable request to the authors.

## Supplementary Materials

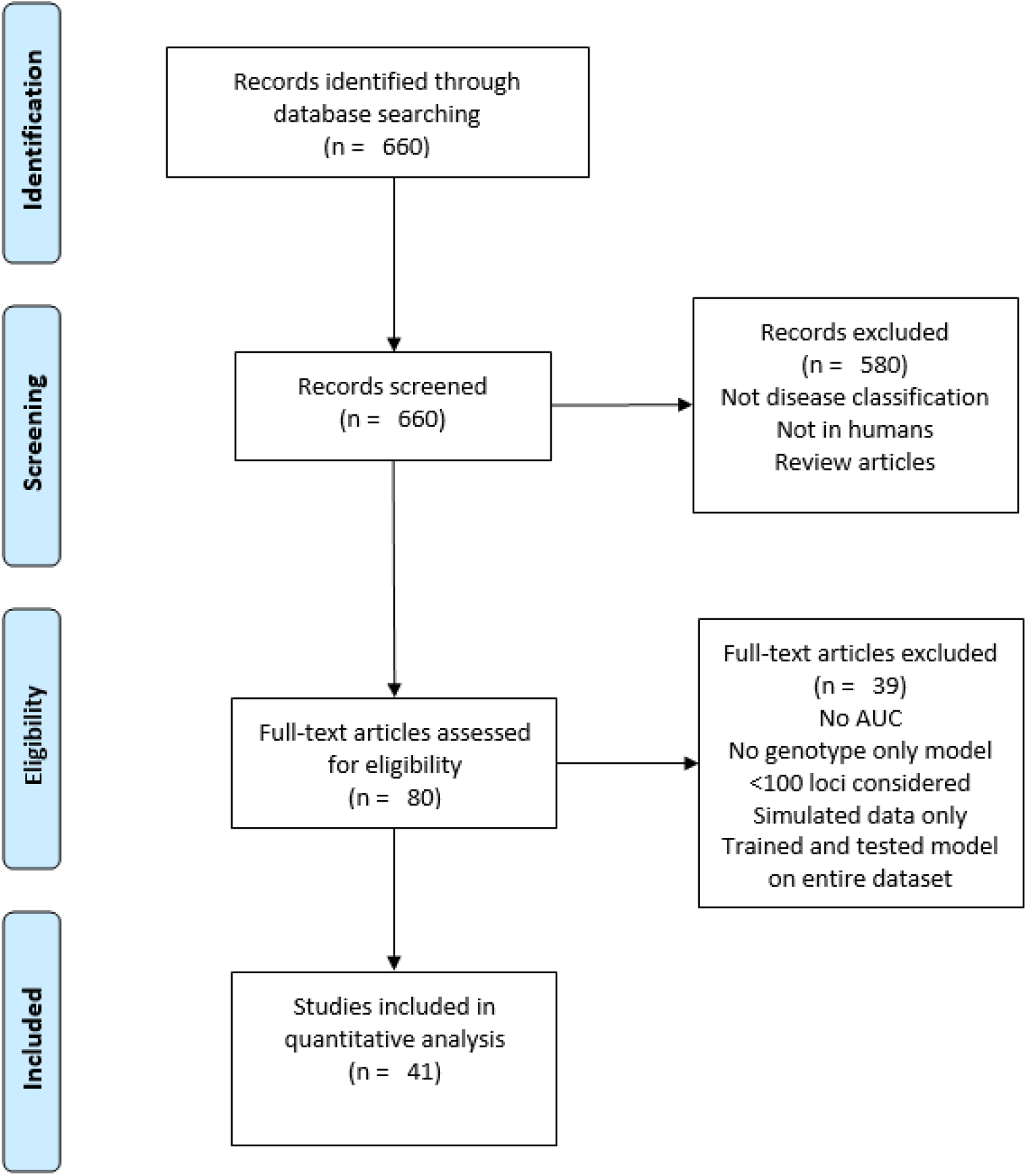

